# Effective real-time transmission estimations incorporating population viral load distributions amid SARS-CoV-2 variants and pre-existing immunity

**DOI:** 10.1101/2024.05.27.24307968

**Authors:** Yu Meng, Yun Lin, Weijia Xiong, Eric H. Y. Lau, Faith Ho, Jessica Y. Wong, Peng Wu, Tim K. Tsang, Benjamin J. Cowling, Bingyi Yang

## Abstract

**Background:** Population-level viral load distribution, measured by cycle threshold (Ct), has been demonstrated to enable real-time estimation of R_t_ for SARS-CoV-2 ancestral strain. Generalisability of the framework under different circulating variants and pre-existing immunity remains unclear.

**Aim:** This study aimed to examine the impact of evolving variants and population immunity on the generalizability of Ct-based transmission estimation framework.

**Methods:** We obtained the first Ct record of local COVID-19 cases from July 2020 to January 2023 in Hong Kong. We modeled the association between daily viral load distribution and the conventional estimates of R_t_ based on case count. We trained the model using data from wave 3 (i.e., ancestral strain with minimal population immunity) and predicted R_t_ for wave 5, 6 and 7 (i.e., omicron subvariants with > 70% vaccine coverage). Cross-validation was performed by training on the other 4 waves. Stratification analysis by disease severity was conducted to evaluate the impact of the changing severity profiles.

**Results:** Trained with the ancestral dominated wave 3, our model provided accurate estimation of R_t_, with the area under the ROC curve of 0.98 (95% confidence interval: 0.96, 1.00), 0.62 (95% CI: 0.53, 0.70) and 0.80 (95% CI: 0.73, 0.88) for three omicron dominated waves 5 to 7, respectively. Models trained on the other four waves also had high accuracy. Stratification analysis suggested potential impact of case severity on model estimation, which coincided with the fluctuation of sampling delay.

**Discussion:** Our findings suggested that incorporating population viral shedding can provide accurate real-time estimation of transmission with evolving variants and population immunity. Application of the model needs to account for sampling delay.

## Introduction

Tracking community transmission in real-time is critical but suffers delays due to the unavoidable right-censoring (i.e., incubation period and delay in case identification) in the conventional methods.^1,2^ Previous work established a novel temporal association between epidemic dynamics and population viral load distribution, measured by cycle threshold (Ct) values from reverse transcription quantitative polymerase chain reaction (RT-qPCR).^3^ We have since applied a simplified method to incorporate the temporal population Ct distribution into real-time transmission estimation, measured by the effective reproductive number R.^4^ While these studies have significantly advanced our understanding of the association between population viral shedding and transmission, they were performed during early waves of the COVID-19 pandemic, and did not account for the viral evolution and immunity derived from natural infections and/or vaccinations.^5^ The generalizability of the identified association to epidemics remained under-investigated, especially in the context of the emerging SARS-CoV-2 variants (e.g., Omicron) and increasing pre-existing immunity, which were found to be associated with shorter duration in viral shedding clearance at individual level.^6^ Additionally, large epidemics with exponential increase in cases could soon exceed testing and surveillance capacity and may further prolong the delays between infection and being diagnosed due to the constrained resources, making it more challenging to derive timely and reliable R_t_ estimates using conventional incidence-based approaches.^1,7^

Here, we examined the impact of the evolving SARS-CoV-2 variants and population immunity on the application of population viral load distribution to estimate transmission, using data of laboratory-confirmed COVID-19 cases from July 2020 to January 2023 in Hong Kong. During this period, the dominant strains transitioned from ancestral strain to Omicron subvariants, and the population shifted from predominantly native to possessing high pre-existing hybrid immunity due to natural infection and vaccinations.^8^ Thus we can examine whether overall association between population-level viral loads and transmission dynamics remained consistent and validate the Ct-based R_t_ estimation method.

## Methods

### Data source

Viral loads of COVID-19 cases were measured as cycle threshold ^9^ values (derived from SARS-CoV-2 RT-qPCR assays targeting E gene) from upper respiratory tract samples, given that they are inversely correlated (i.e. lower Ct values imply higher viral loads).^10,11^ We collected the clinical, epidemiological and demographic data of each local cases from the Hospital Authority (HA) and the Department of Health of the Government of Hong Kong during the observation period, including their first recorded Ct values, date of sampling, clinical outcomes and date of symptom onset. We also classified cases as mild-to-moderate, serious, critical and fatal according to their clinical outcomes.^8,12^ In this study, we constrained analyses to two severity groups, namely mild-to-moderate and severe groups (combining serious, critical and fatal cases into a single group).

## Statistical Analysis

### R_t_ estimation based on case counts (incidence-based R_t_)

R_t_ estimation on COVID-19 should be based on infection time because of its pre-symptomatic transmission^1,7^. Thus, we applied robust incidence deconvolution estimator ^13^ with delay from infection to reporting, to reconstruct the epidemic curve by infection time. Then we estimate the incidence-based R based on daily local cases numbers using Cori’s method.^7^ In this framework, the incidence-based R_t_ was the ratio of the number of new cases to the total infectiousness of infective individuals, which was the convolution of the incubation period and the infectiousness relative to onset.^14^ We conducted inference by a Markov chain Monte Carlo algorithm to estimate R.^15,16^ More details about incidence-based R estimation was described elsewhere.^1,4^

### Incorporating Ct distribution into R_t_ estimates (Ct-based R_t_)

Hong Kong experienced multiple epidemic waves between 1 January 2020 and 29 January 2023, and it was dominated by local transmissions from wave 3.^8^ We analyzed the first record for confirmed local COVID-19 cases (i.e., no travel outside Hong Kong during incubation) with available Ct values. The whole observed period was split into three uninterrupted sub-periods, i.e., 1 July to 31 August 2020 (wave 3), 1 November 2020 to 31 March 2021 (wave 4) and 1 January 2022 to 29 January 2023 (wave 5 and 6), to fit generalized additive model (GAM) to characterize the population distribution of viral load separately (Appendix).

To assess the relationship between population-level distribution of viral loads and incidence-based R_t_, we first calculated the Spearman’s rank correlation coefficient (ρ) between daily Ct distribution (i.e., mean and skewness) and the natural log-transformed incidence-based R_t_ (Table 1). We fitted a linear regression model of the daily mean and skewness of Ct on log-transformed incidence-based R (established from our previous study^4^), using training data from the third wave (i.e., ancestral strain wave). We applied the fitted model to predict R_t_ using population-level Ct distribution in the fourth to sixth waves (i.e., ancestral strain and Omicron variants waves), respectively. We included a 31-day period in the training set (e.g., 19 July to 18 October 2020 for wave 3), consisting of 10 days before and 20 days after the day when local cases peaked in that wave, as suggested by previous study.^4^ We determined the case peak by computing the 5-day rolling average of confirmed local cases, to minimize the impact of sudden reporting changes.

**Table 1.** Spearman’s correlation coefficients (ρ) between Ct and the natural log-transformed incidence-based *Rt*.

|  | Wave 3<br>(Jul - Aug 2020)<sup>^</sup> |  | Wave 4<br>(Nov 2020 - Mar 2021) |  | Wave 5<br>(Jan 2022 - May 2022) |  | Wave 6<br>(May 2022 - Sep 2022) |  | Wave 7<br>(Oct 2022 - Jan 2023) |  |
| --- | --- | --- | --- | --- | --- | --- | --- | --- | --- | --- |
| | $\rho$ | *P*-value<sup>^</sup> | $\rho$ | *P*-value<sup>^</sup> | $\rho$ | *P*-value<sup>^</sup> | $\rho$ | *P*-value<sup>^</sup> | $\rho$ | *P*-value<sup>^</sup> |
| **Ct mean** | -0.79 | <0.001 | -0.51 | <0.001 | -0.69 | <0.001 | -0.14 | 0.105 | -0.58 | <0.001 |
| **Ct skewness** | 0.8 | <0.001 | 0.27 | 0.001 | 0.62 | <0.001 | 0.11 | 0.221 | 0.55 | <0.001 |

<sup>^</sup>Two-sided P-values that were rounded to 3 decimal places.

We evaluated the model prediction using the area under the receiver operating characteristic curve (AUC) as the primary metric,^17,18^ fitted to the binary outcome of whether Ct-based (predicted) and incidence-based (observed) R_t_ above 1. This threshold was chosen because an R_t_ above 1 indicates a growing epidemic trend, while values below 1 indicate a decreasing trend, which plays a crucial role in providing early warnings for public health. ^19^ We also computed the directional consistency as the proportion of days during the prediction period when the predicted and observed R were either simultaneously below or exceeding 1.^4^

### Cross-validation between epidemic waves

To reflect the government’s relaxation of entry restrictions and the introduction of new strains, such as XBD, BF.7 and BQ.1.1, wave 6 was further split into waves 6 (23 May to 30 September 2022) and 7 (1 October 2022 to 29 January 2023).^8,20^ Then, we trained the model on data from wave 4 to 7, separately, to evaluate the generalizability of our method. As described above, we included a 31-day training period of wave 4 (24 November to 24 December 2020), wave 5 (21 February to 23 March 2022), wave 6 (23 August to 22 September 2022) and wave 7 (19 December 2022 to 18 January 2023), respectively. With each training set, the other waves as well as wave 3 were used as test sets. We evaluated the model predictions using AUC and directional consistency. We excluded from wave 5 the forecasts made between 1 January and 6 February 2022 due to the huge fluctuation of incidence-based R_t_.

### Stratified analysis of two symptom severity groups

To further validate the Ct-based framework, we assess the impact of the changing severity profiles in confirmed COVID-19 cases. This involved characterizing the temporal changes of first Ct distribution and delay of onset to sampling stratified by the retrospectively classified clinical severity. In particular, the retrospective subsettings of severity were determined based on their clinical outcomes that have already occurred, which may have been unknown during the collection of the initial Ct values. Thus, we also characterized the distribution of the delays between the initial records and clinical outcomes. Subsequently, we fitted the established model from Ct data from either mild-to-moderate or severe group only in wave 3, 4, 5, 6, and 7, respectively, and then used model and date from corresponding severity group to estimate the scenarios in wave 5, 6 and 7.

All statistical analyses were conducted in R version 4.3.1 software (R Foundation for Statistical Computing, Vienna, Austria).

## Results

### COVID-19 waves in Hong Kong

COVID-19 cases were detected from people with respiratory symptoms or high risk of exposures (e.g., close contacts and occupational exposure) and confirmed with RT-qPCR between January 2020 and 6 February 2022,^4^ covering wave 3, 4 and early wave 5. Contact tracing were suspended after 7 February 2022 (waves 5 and 6), and self-reported positive rapid antigen test (RAT) were also recorded as cases after 26 February 2022.^21^ A decline in the incidence-based R was observed throughout April and May 2022 before cases number increased again in June partially due to the emergence of the Omicron BA.4/BA.5 which grew steadily and eventually replaced BA.2 as the dominant variants, with BA.5 having an absolute advantage since later August 2022.^22^ By later January 2023, sublinages BA.2 and BA.5 became the dominating lineages in Hong Kong.^23^ Two COVID-19 vaccines (CoronaVac [Sinovac] and BNT162b2 [BioNTech/Fosun Pharma/Pfizer]) were provided for free in Hong Kong government since February 2021 (during and after wave 4).^24^ As of January 2, 2022 (wave 5), approximately 66% of the population had received at least one vaccine dose.^25^

We included local cases from July 2020 to January 2023, covering wave 3 (1 July to 31 August 2020) and wave 4 (1 November 2020 to 31 March 2021) caused by the ancestral strain, while wave 5 (1 January to 22 May 2022), wave 6 and 7 (23 May 2022 to 29 January 2023) were caused by Omicron BA.2 and BA.4/BA.5, respectively (Figure).^8,12^

In total 2,790,814 local RT-qPCR confirmed COVID-19 cases and self-reported RAT positives were recorded through wave 3 and 7, with about 43%, 19% and 38% of the cases recorded in wave 5, 6 and 7, respectively (Table S2). Ct values were available for 114,714 cases included, with 95% (n=3,043), 96% (n=5,225), 4% (n=51,372), 4% (n=21,835), and 3% (n=33,239) in waves 3 to 7, respectively.

**Figure.**
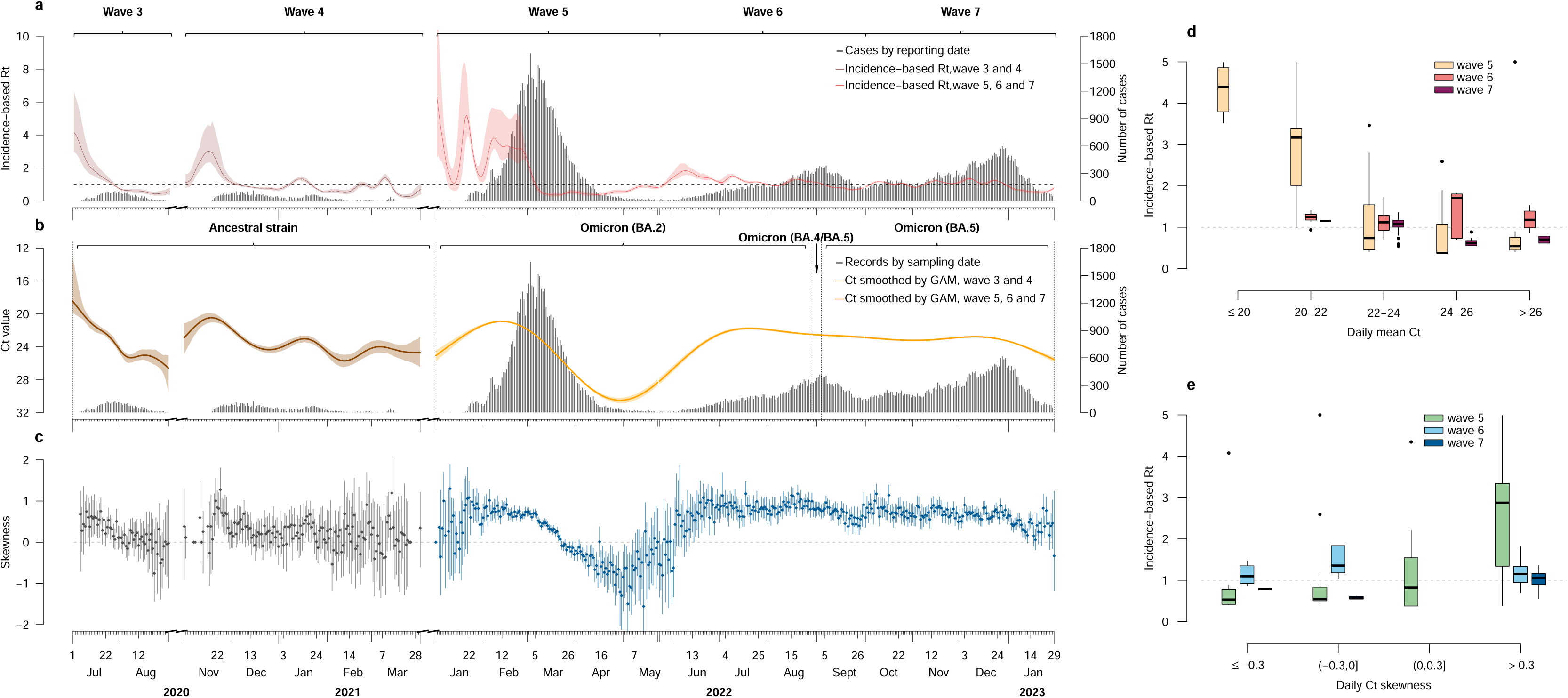
Temporal distribution of incidence-based R_t_ (estimated by cases count), and population-level Ct values (measured by daily mean and skewness). **a**. Locally confirmed COVID-19 cases with available Ct values by date of reporting and incidence-based R_t_ estimated by cases count. Black bars indicate daily case counts. Lines and shaded areas indicate the means and 95% credible intervals (CrIs) for incidence-based R_t_ over the entire observed period. **b.** Temporal distribution of population-level Ct values and main dominant strains. Black bars indicate the number of daily collected samples. Brown lines and shaded areas indicate the average and 95% confidence intervals (CIs) of Ct values estimated by a generalized additive model (GAM) during the third and fourth waves which were dominated by ancestral strain. Orange lines and shaded areas correspond to the fifth and sixth waves which were dominated by Omicron variants. **c.** Temporal distribution of Ct skewness. Dots and vertical lines represent the mean and 95% CIs of daily Ct skewness. **d, e.** Correlations between the incidence-based R_t_ and Ct mean (panel d) or skewness (panel e) during the fifth and sixth waves. Box plots indicate the interquartile ranges (IQR) and medians of the incidence-based R_t_ under various intervals of daily Ct mean (panel d) and skewness (panel e).

### Correlations between population-level Ct distribution and incidence-based *H_i_*

We examined the correlation between the temporal population-level distribution of Ct values (measured by daily mean and skewness) and the local transmission dynamics (measured by the incidence-based R_t_). We observed consistent temporal associations between population Ct distribution and incidence-based R_t_ in the Omicron variants-dominated waves 5 and 7, as seen for the ancestral strain-dominated waves 3 and 4 (Figure). Higher incidence-based R_t_ were observed with lower average Ct values (Spearman’s correlation coefficient, ρ = –0.69, *P* < 0.001 for wave 5 and ρ = –0.58, *P* < 0.001 for wave 7) and as Ct skewed towards lower values (ρ = 0.62, *P* < 0.001 for wave 5 and ρ = 0.55, *P* < 0.001 for wave 7) (Table 1), although such relationships were not significant for wave 6 (Table 1).

### Estimating Ct-based H_i_ for waves with changing dominant variant and population immunity

We evaluated the performance of our approaches in later waves to examine the impact of changing dominating SARS-CoV-2 variants (wave 4-7) and population immunity (wave 5-7) on the Ct-based real-time COVID-19 transmission. We trained the model using the 31-day data around peak of the ancestral strain-dominated wave 3 (as suggested previously ^4^) and applied the trained model to predict R_t_ using daily Ct distributions in waves 4 to 7 (i.e., testing periods), separately. We found that the Ct-based method provided accurate real-time estimations of R_t_ during testing periods (Table 2), with area under the receiver operating characteristic (ROC) curve (AUC) of 0.68 (95% confidence interval (CI), 0.60, 0.75) (test set, wave 4), 0.98 (95% CI, 0.96, 1) (test set, wave 5), 0.62 (95% CI, 0.53, 0.70) (test set, wave 6) and 0.80 (95% CI, 0.73, 0.88) (test set, wave 7), respectively (Table 2).

**Table 2.** Model performance using different training periods to estimate Ct-based *Rt* in the other four waves.

|  | Training period: wave 3<sup>*</sup><br>( 19 Jul-18 Aug 2020) |  | Training period: wave 4<br>(24 Nov - 24 Dec 2020) |  | Training period: wave 5<br>(21 Feb -23 Mar 2022) |  | Training period: wave 6<br>(23 Aug-22 Sep 2022) |  | Training period: wave 7<br>(19 Dec 2022-18 Jan 2023) |  |
| --- | --- | --- | --- | --- | --- | --- | --- | --- | --- | --- |
|  | AUC<sup>a</sup> | Consistency<sup>b</sup> | AUC<sup>a</sup> | Consistency<sup>b</sup> | AUC<sup>a</sup> | Consistency<sup>b</sup> | AUC<sup>a</sup> | Consistency<sup>b</sup> | AUC<sup>a</sup> | Consistency<sup>b</sup> |
| **Wave 3** | 0.94<br>(0.86, 1.00) | 94.7% | 0.92<br>(0.84, 1.00) | 94.7% | 0.80<br>(0.70, 0.90) | 77.2% | 0.91<br>(0.83, 1.00) | 93.0% | 0.92<br>(0.85, 1.00) | 93.0% |
| **Wave 4** | 0.68<br>(0.60, 0.75) | 71.9% | 0.69<br>(0.61, 0.76) | 73.3% | 0.67<br>(0.59, 0.75) | 66.4% | 0.70<br>(0.62, 0.77) | 74.7% | 0.71<br>(0.64, 0.79) | 74.7% |
| **Wave 5** | 0.98<br>(0.96, 1.00) | 97.1% | 0.98<br>(0.96, 1.00) | 97.1% | 0.96<br>(0.92, 1.00) | 96.2% | 0.99<br>(0.97, 1.00) | 98.1% | 0.98<br>(0.96, 1.00) | 97.1% |
| **Wave 6** | 0.62<br>(0.53, 0.70) | 67.9% | 0.62<br>(0.53, 0.70) | 67.9% | 0.53<br>(0.49, 0.57) | 35.9% | 0.66<br>(0.58, 0.74) | 59.5% | 0.66<br>(0.57, 0.75) | 68.7% |
| **Wave 7** | 0.80<br>(0.73, 0.88) | 80.2% | 0.81<br>(0.73, 0.88) | 81.0% | 0.49<br>(0.46, 0.53) | 37.2% | 0.53<br>(0.50, 0.56) | 41.3% | 0.67<br>(0.59, 0.75) | 62.8% |
| **Overall<sup>c</sup>** | 0.78<br>(0.74, 0.82) | 78.1% | 0.83<br>(0.79, 0.86) | 82.9% | 0.53<br>(0.49, 0.57) | 51.2% | 0.69<br>(0.65, 0.73) | 73.4% | 0.80<br>(0.76, 0.84) | 80.6% |

Incidence-based $R_t$ was natural log-transformed. <sup>\*</sup> The main model used to estimate Ct-based $R_t$
a AUC: area under the receiver operating characteristic curve (ROC)
b Directional consistency.
c Overall: combined all test sets into one to calculate corresponding AUC and directional consistency

We validated our methods by training the model using data from waves 4 to 7. The model demonstrated high accuracy in predicted R_t_, when using wave 3 and 5 as test sets (Table 2). For example, AUC of predicted Ct-based R_t_ for Omicron variants-dominated wave 5 was 0.98 (95% CI, 0.96, 1) (trained with wave 4), 0.99 (95% CI, 0.97, 1) (trained with wave 6) and 0.98 (95% CI, 0.96, 1) (trained with wave 7). Similarly, retrospective R_t_ estimates for ancestral strain-dominated wave 3 indicated robust accuracies with AUC of 0.92 (95% CI, 0.84, 1) (trained with wave 4), 0.80 (95% CI, 0.70, 0.90) (trained with wave 5), 0.91 (95% CI, 0.83, 1) (trained with wave 6) and 0.92 (95% CI, 0.85, 1) (trained with wave 7). However, despite accurate prediction of wave 5, utilizing it as a training set led to suboptimal performance, as evidenced by the AUC of predicted Ct-based Rt for wave 6 (0.53, 95% CI, 0.49, 0.57) and wave 7 (0.49, 95% CI, 0.46, 0.53). Of note, Ct-based estimates on wave 6 were generally weaker than other waves, with AUC equal to 0.62 (95% CI, 0.53,0.70) (trained with wave 3), 0.62 (95% CI, 0.53, 0.70) (trained with wave 4), 0.53 (95% CI, 0.49, 0.57) (trained with wave 5), 0.66 (95% CI, 0.57, 0.75) (trained with wave 7).

### The impact of severity on the association between population-level Ct distribution and incidence-based ***H_i_***

There were more severe (serious, critical, and fatal cases) cases recorded during Omicron waves compared with ancestral strain waves. 40% cases were retrospectively classified as severe in wave 5 and 7, compared with less than 25% in wave 3 and 4 (Table S3). As suggested, we retrospectively assess Ct-based Rt estimates based on subsets of Ct values from cases with two distinct degrees of severity, including mild-to-moderate group and severe group, to further validate the generalizability of our model. A slight delay was found in the temporal distribution of Ct values between mild-to-moderate and severe cases during wave 5 and 6 (Figure S2), which coincided with the different delays of onset to sampling between mild-to-moderate and severe group during February 2022 (wave 5), April 2022 (wave 5), and August 2022 (wave 6).

Results for the model trained in wave 3 suggested that using Ct values from mild-to-moderate or severe only group yielded decreased AUCs (e.g., 0.52, 95% CI, 0.48, 0.56 for wave 5, using severe cases only) compared to using data from all cases (e.g., 0.98, 95%CI, 0.96, 1 for wave 5, using all cases), and the model trained in wave 4 show similar trend (Table S4). In contrast, for the training period with increasing proportion of severe cases (Table S3), using Ct from one severity group could yield higher AUCs compared to using Ct from all cases. For instance, the AUC of Ct-based Rt for wave 6 was 0.63 (95% CI, 0.55, 0.71) (trained with mild-to-moderate cases from wave 5), compared to 0.53 (95% CI, 0.49, 0.57) (trained with all cases from wave 5) (Table S4).

## Discussion

In this study, we demonstrated that the association between population viral load distribution and epidemics, derived from ancestral strain with minimal population immunity, remained highly informative during the subsequent SARS-CoV-2 epidemic waves, including those dominated by the Omicron subvariants in populations with significant pre-existing immunity. Our findings also suggested several circumstances in which the model predictions may be conservative, such as amid fluctuations in sample representativeness or during plateaued local epidemics, thereby providing insights into the broader applicability of the model.

Reduced viral shedding durations were observed for vaccinated individuals and those infected with the Omicron variant; however, disparities in viral loads were minimal during the early stages of infection.^6^ Therefore, our approach employing the initial Ct post-confirmation may be minimally affected by variants and vaccinations, consistent with previous studies measuring the transmission using various measurements of transmission (e.g., growth rate).^5,26–28^

For surveillance based primarily on symptoms and contact tracing,^4^ Ct samples could be delays in case identification or sample collection, resulting in an inaccurate reflection of the overall viral load in the population. For example, we observed lower population Ct values for severe cases in early wave 5 but higher values in late wave 5 and early wave 6, compared to mild-to-moderate cases (Figure S2), coincided with observed disparities in delays from symptom onset to sample collection (Figure S8). Our stratified analyses of sample severity profile also revealed a complex impact of severity on the Ct-based estimation of Rt, highlighting the importance of stable sample representativeness. During waves 3 and 4 in Hong Kong, when intensive surveillance and contact-tracing adopted amid low virus circulation in the community, models trained on all cases outperformed those trained only on mild-to-moderate cases, as they better represented the full spectrum of case exposure time distribution (Figure S3-S7). Conversely, when training the model using data that has significant increases in the proportion of severe samples with delayed reporting, the model may inaccurately associate these changes in severity profile and reporting to changes in transmission, leading to reduced model performance (Table S4). This observation is consistent with the shorter delays observed between initial Ct reports and clinical outcomes during Omicron waves compared to the delays observed during waves 3 and 4 (Figure S9). Future research on the impact surveillance delays and sample representativeness on association between population viral shedding and transmission would better inform the applicability of the Ct-based estimation framework.

The accuracy of Ct-based R_t_ estimates for wave 6 was not optimal, irrespective of whether the model was trained with the data from the same wave or other waves, with deviations mainly occurring in late May and early June 2022 (Figure S1). A possible reason could be the low and relatively consistent community transmission, as evidenced by the incidence-based Rt stably fluctuating around 1 and its Gini coefficient of 0.140 (95% CI, 0.125, 0.156) (Table S5). Simultaneously, fewer samples available to obtain Ct values led to increased uncertainties surrounding the Rt estimates, as suggested by our previous study.^4^ Whilst increased AUCs for testing sets in wave 4 and 6 were observed when we excluded days with less than 30 or 60 records (Table S6). Besides, the training period included for wave 6 coincided with the presence of multiple Omicron variants, including BA.2 and BA.4/BA.5. These variants were indicated to exhibit distinct viral kinetics and infectiousness,^29,30^ however, limited genotyping data pose challenges in evaluating the impact of the transition period when competing Omicron variant coexisted on our model’s performance. Future study on model predictions during co-circulation or transition period are warranted for better understanding of our model applications under these complex situations.

Our work has several limitations. Firstly, we were unable to further examine the effect of vaccines and variants due to the limited individual data. Secondly, the reduction in case ascertainment as non-pharmaceutical interventions were relaxed^8^ and the potential bias towards reporting severe cases in the later epidemic waves may have affected the accuracy of R*t* estimation based on case counts. Further research is needed to address this issue and refine the model accordingly. Additionally, it is worth noting that our model requiring less computation efforts and crude metric AUC enables robust binomial estimation of R_t_ values exceeding 1 or descending below 1. Whilst it may not afford absolute quantitative estimates, as was demonstrated in the nowcast for wave 5 (Table S7 and Figure S1-C). Therefore, it is necessary to validate the model in other populations to improve its generalizability.

Our study provides valuable insights into the potential of population-level Ct distribution as a predictive tool for timely assessing Rt during waves characterized by variants dominating and population immunity shifting. These findings suggest the potential generalizability of this simplified framework across various settings and situations. It is important to exercise caution when interpreting the results due to the fluctuation of sampling delay and severity proportion. Further research is required to validate these findings and address the issue of estimating Rt when daily records are limited and when community transmission were stable and consistent.

## Supporting information

Figures for supplemental materials

## Data Availability

All data produced in the present study are available upon reasonable request to the authors

## Acknowledgements

We thank the Department of Health and Hospital Authority of the Food and Health Bureau of the Government of Hong Kong for providing the data for the analysis. This project was supported by the Health and Medical Research Fund, Food and Health Bureau, Government of the Hong Kong Special Administrative Region (grant no. 22210552, B.Y.), the Theme-based Research Scheme (Project No. T11-705/21-N; B.J.C.) and the general research fund (Project No. 17100822; T.K.T.) of the Research Grants Council of the Hong Kong SAR Government. We also thank Justin K. Cheung and Chloe S. Chui for their help in managing and collecting the data.

## Author contributions

All authors meet the ICMJE criteria for authorship. The study was conceived by B.Y. and B.J.C. Y.M., Y.L., W.X., E.H.Y.L., F.H., J.Y.W., W.P. and T.K.T. prepared the data. B.Y., Y.M. and Y.L. developed the model. Y.M. conducted the data analyses. Y.M., B.Y., T.K.T., and B.J.C. interpreted the results. Y.M. wrote the first draft of the paper. All authors provided critical review and revision of the text and approved the final version.

## Competing interests

B.J.C. consults for AstraZeneca, Fosun Pharma, GSK, Haleon, Moderna, Roche, Sanofi Pasteur, and Pfizer. The remaining authors declare no competing interests.

## Notes

### Funding Statement

This study was funded by the Health and Medical Research Fund, Food and Health Bureau, Government of the Hong Kong Special Administrative Region (grant no. 22210552, B.Y.), the Theme-based Research Scheme (Project No. T11-705/21-N; B.J.C.) and the general research fund (Project No. 17100822; T.K.T.) of the Research Grants Council of the Hong Kong SAR Government.

### Author Declarations

Ethical approval for this study was obtained from the Institutional Review Board of The University of Hong Kong/Hospital Authority Hong Kong West Cluster.

