## Supplemental materials for "Effective real-time transmission estimations incorporating population viral load distributions amid SARS-CoV-2 variants and pre-existing immunity"

**Supplementary Methods**

*1. Estimation of Rt based on case counts (incidence-based Rt)*

We estimated the incidence-based $R_{t}$for local COVID-19 cases based on Cori’s method and robust incidence deconvolution estimator ^1-3^. The deconvolution approach in Becker et al.^4^ was used to obtain the actual (unobserved) number of new cases confirmed at day *t*, $Y\left( t \right)$, from the epidemic curve by onset time and a given distribution for delay from infection to report. While the latter was obtained by convolving incubation period (mean 5.2 days, SD 3.9 days)^5^ and an empirical distribution of delay from onset to report (mean 4.7 days, SD 3.2 days). For the transmission, we modeled a Poisson process, namely

$$Y\left( t \right) \sim Poisson\left\{ R_{t}\sum_{k=1}^{t-1} Y(k)\omega_{L}(t-k) \right\}$$

Hence, the daily local$R_{t}$ (i.e., the incidence-based $R_{t}$) was the ratio of the number of new cases, $Y\left( t \right)$, to the total infectiousness of infective individuals at time t, given by $\sum_{k=1}^{t-1} Y(k)\omega_{L}(t-k)$, where $\omega_{L}(t-k)$ (the probability distribution of being infectious$t-k$ days after infection) was estimated by convolution of the incubation period and the infectiousness relative to onset^6^. To enhance interpretation, we applied the smoothing method in Cori et al.^3^ and a time window of size equal to 14 to assume a constant transmissibility and to avoid instability. We also used a Markov chain Monte Carlo algorithm to update the model parameter for $R_{t}$estimation^7^. To account for the uncertainty in input parameter, such as incubation period and infectiousness distribution, we performed bootstrap approach described in Salje et al.^8^ to reconstruct 200 epidemic curves and finally obtained the mean, 2.5% and 97.5% quantiles for 200 $R_{t}$estimates at each time point (Figure a). More details about incidence-based $R_{t}$estimation was described elsewhere1,8.

*2. Characterizing the temporal distribution of population-level viral loads*

We characterized the temporal distribution of population-level viral loads by mean ($\bar{\chi}_{t}$) and skewness ($b_{t}$) on a given day, $t$:

$$\bar{\chi}_{t} = \frac{1}{n_{t}} \sum_{i=1}^{n_{t}} y_{t,i}$$

$$b_{t} = \frac{\frac{1}{n_{t}}\sum_{i=1}^{n_{t}} {(y_{t,i} - \bar{\chi}_{t})}^{3}}{{[\frac{1}{n_{t}-1}\sum_{i=1}^{n_{t}} {(y_{t,i}- \bar{\chi}_{t})}^{2}]}^{\frac{3}{2}}}$$

Where $y_{t,i}$represented the $i$th ($i$ = 1, 2, …, $n_{t}$) of the total $n_{t}$ Ct values that were sampled on day$t$. 95% CIs of the daily skewness $b_{t}$ were calculated from 500 bootstraps (Figure c).

To characterize the temporal distribution of Ct values, generalized additive model (GAM) was fitted over the 3 study timelines separately.

$$y_{j,t}=\alpha_{0}+s\left（ t \right）$$

where $y_{j,t}$ was the first recorded Ct for local case $j$ (*t* is the calendar date when the sample collected), and $s\left（ t \right）$ was the smooth function for a given date *t*. 95% confidence intervals (CIs) of the smoothed average daily Ct were derived from 500 bootstraps (Figure a). In each bootstrap, we resampled from the data on cases’ first available Ct values and refitted the GAM. We imputed the daily Ct distributions using the average of that within the preceding 7 days when no samples were collected on that day. Additionally, we calculated Gini coefficients for incidence-based 𝑅𝑡 and population Ct to assess their distribution balance.

*3. Main model*

We fitted a log-linear regression model of the daily mean and skewness of Ct on incidence-based $R_{t}$ (main model obtained from our previous study), using 31-day data from the third wave (i.e., ancestral strain), and used it to predict 𝑅_𝑡_ in the fourth (i.e., ancestral strain) to sixth waves (i.e., Omicron variants), respectively.

$$\ln\left( R_{t} \right)= y_{0}+y_{\bar{\chi}}\bar{\chi}_{t}+y_{b}b_{t}$$

where $y_{\bar{\chi}}$ and $y_{b}$were coefficients for daily mean and skewness of Ct values from the regression model. The coefficients from the regression model were reported in Supplementary Table S1 after exponential transformation.

*4. Case classification based on the severity statues*

Our previous study divided confirmed cases into four groups according to their clinical manifestations, namely mild-to-moderate, serious, critical and fatal cases.^9^ In this study, we combined the serious, critical and fatal cases into one group (severe group), and the classification criteria were as follow:

1) For mild-to-moderate cases, those who were defined as neither serious nor fatal were then classified as mild-to-moderate cases.

2) For serious cases, patients who meet one of the following criteria:

a. treated with dexamethasone and/or baricitinib (or IV tocilizumab);

b. experienced O2 desaturated;

c. received oxygen supplement of 3 liters per minute or more.

3) For critical cases, patients who meet one of the following criteria:

a. admitted to ICU;

d. required intubation or extracorporeal membrane oxygenation (ECMO);

c. in shock.

4) For fatal cases, patients who died within 28 days of their first positive test for SARS-CoV-2 were classified as fatal.

*5. Sensitivity analysis of sample counts*

For the waves in which incidence-based 𝑅𝑡 remained relatively stable and yielded suboptimal estimates, we performed sensitivity analyses by excluding days with less than 30 or 60 Ct records in the testing sets to re-calculate AUC and directional consistency. This allowed us to further evaluate the performance of our simplified Ct-based framework, and the results were summarized in Table S6.

**Supplementary Tables**

| **Table S1.** Associations between population Ct distributions and incidence-based 𝑅𝑡. Regression coefficients (*β)*, their 95% confidence intervals (CIs) and P-values were computed from the main model. | | | |
| --- | --- | --- | --- |
|  | *β* (95%CIs) | P-value^^^ | Adjusted R square^*^ |
| **Main model (training period between 19 Jul to 18 Aug 2020)** | | | |
| **Ct mean** | 0.86(0.8,0.93) | <0.001 | 0.69 |
| **Ct skewness** | 1.27(0.75,2.15) | 0.359 |  |
| **Validation model 1 (training period between 24 Nov to 24 Dec 2020)** | | | |
| **Ct mean** | 0.9(0.87,0.93) | <0.001 | 0.8 |
| **Ct skewness** | 1.26(1.04,1.53) | 0.022 |  |
| **Validation model 2 (training period between 21 Feb to 23 Mar 2022)** | | | |
| **Ct mean** | 0.37(0.2,0.68) | 0.002 | 0.71 |
| **Ct skewness** | 0.03(0,1.51) | 0.078 |  |
| **Validation model 3 (training period between 23 Aug to 22 Sep 2022)** | | | |
| **Ct mean** | 0.81(0.75,0.87) | <0.001 | 0.78 |
| **Ct skewness** | 0.98(0.65,1.46) | 0.906 |  |
| **Validation model 4 (training period between 19 Dec 2022 to 18 Jan 2023)** | | | |
| **Ct mean** | 0.77(0.69,0.85) | <0.001 | 0.8 |
| **Ct skewness** | 0.88(0.48,1.63) | 0.674 |  |
| ^*^Adjusted R square of the corresponding model. |  |  |  |
| ^^^ Two-side P-values that were derived from t-tests for whether the coefficient was significantly different from 0, and were rounded to 3 decimal places. | | | |

| **Table S2**. Number of local cases confirmed by RT-qPCR and RAT in Hong Kong during the study period. | | | | | |
| --- | --- | --- | --- | --- | --- |
|  | **Days interval** | **RAT-positive** | **PCR-positive** | **Total confirmed cases** | **Cases with Ct values**^*^ |
| **Wave 3** | 62 | - | 3,217 (100%) | 3,217 | 3,043 (95%) |
| **Wave 4** | 151 | - | 5,426 (100%) | 5,426 | 5,225 (96%) |
| **Wave 5** | 142 | 448,957 (38%) | 746,829 (62%) | 1,195,786 | 51,372 (4%) |
| **Wave 6** | 131 | 362,723 (68%) | 170,413 (32%) | 533,136 | 21,835 (4%) |
| **Wave 7** | 121 | 834,144 (79%) | 219,095 (21%) | 1,053,249 | 33,239 (3%) |
| **Total** | 607 | 1,645,824 (59%) | 1,144,980 (41%) | 2,790,814 | 114,714 (4%) |
| ^*^ The denominator is the total number of local confirmed cases. | | | | | |

| **Table S3**. Number of local cases categorized by different symptom severity levels in Hong Kong during the study period. | | | | | |
| --- | --- | --- | --- | --- | --- |
| **Case classification** | **Wave 3** | **Wave 4** | **Wave 5** | **Wave 6** | **Wave 7** |
| **Mild-to-moderate** | 2372 (77.9%) | 4,088 (78.2%) | 30,871 (60.1%) | 16,358 (74.9%) | 20,174 (60.7%) |
| **Serious/critical/fatal** | 671 (22.1%) | 1,137 (21.8%) | 20,501 (39.9%) | 5,477 (25.1%) | 13,065 (39.3%) |
| **Total** | 3,043 | 5,225 | 51,372 | 21,835 | 33,239 |

| **Table S4.** Area under the ROC curve and directional consistency of Ct-based 𝑅𝑡 using records form either mild-to-moderate or severe cases during Omicron Waves in Hong Kong. | | | | | | |
| --- | --- | --- | --- | --- | --- | --- |
|  | **Wave 5** | | **Wave 6** | | **Wave 7** | |
|  | **(Jan 2022 - May 2022)** | | **(May 2022 - Sep 2022)** | | **(Oct 2022 - Jan 2023)** | |
|  | **AUC^a^** | **Consistency^b^** | **AUC^a^** | **Consistency^b^** | **AUC^a^** | **Consistency^b^** |
| **Training period: wave 3 (6 Jul-31 Aug 2020)** | | | | | | |
| **All cases** | 0.98 (0.96, 1.00) | 97.1% | 0.62 (0.53, 0.7) | 67.9% | 0.8 (0.73, 0.88) | 80.2% |
| **Mild-to-moderate** | 0.99 (0.97, 1.00) | 98.1% | 0.56 (0.48, 0.64) | 67.2% | 0.75 (0.67, 0.83) | 79.3% |
| **Severe** | 0.52 (0.48, 0.56) | 76.2% | 0.50 (0.50, 0.50) | 30.5% | 0.50 (0.50, 0.50) | 37.2% |
| **Training period: wave 4 (20 Nov - 19 Dec 2020)** | | | | | | |
| **All cases** | 0.98 (0.96, 1.00) | 97.1% | 0.62 (0.53, 0.70) | 67.9% | 0.81 (0.73, 0.88) | 81.0% |
| **Mild-to-moderate** | 0.99 (0.97, 1.00) | 98.1% | 0.62 (0.53, 0.70) | 70.2% | 0.75 (0.67, 0.83) | 76.9% |
| **Severe** | 0.96 (0.92, 0.99) | 93.3% | 0.55 (0.46, 0.64) | 51.1% | 0.72 (0.65, 0.79) | 66.9% |
| **Training period: wave 5 (21 Feb -22 Mar 2022)** | | | | | | |
| **All cases** | 0.96 (0.92, 1.00) | 96.2% | 0.53 (0.49, 0.57) | 35.9% | 0.49 (0.46, 0.53) | 37.2% |
| **Mild-to-moderate** | 0.98 (0.96, 1.00) | 97.1% | 0.63 (0.55, 0.71) | 56.5% | 0.49 (0.42, 0.57) | 41.3% |
| **Severe** | 0.97 (0.95, 1.00) | 96.2% | 0.57 (0.51, 0.64) | 45.8% | 0.57 (0.52, 0.62) | 47.1% |
| **Training period: wave 6 (28 Aug-26 Sep 2022)** | | | | | | |
| **All cases** | 0.99 (0.97, 1.00) | 98.1% | 0.66 (0.58, 0.74) | 59.5% | 0.53 (0.50, 0.56) | 41.3% |
| **Mild-to-moderate** | 0.90 (0.83, 0.98) | 95.2% | 0.72 (0.65, 0.79) | 64.9% | 0.49 (0.45, 0.54) | 38.0% |
| **Severe** | 0.95 (0.92, 0.98) | 92.4% | 0.58 (0.50, 0.66) | 48.9% | 0.60 (0.55, 0.66) | 51.2% |
| **Training period: wave 7 (18 Dec 2022-16 Jan 2023)** | | | | | | |
| **All cases** | 0.98 (0.96, 1.00) | 97.1% | 0.66 (0.57, 0.75) | 68.7% | 0.67 (0.59, 0.75) | 62.8% |
| **Mild-to-moderate** | 0.97 (0.93, 1.00) | 98.1% | 0.65 (0.56, 0.74) | 67.9% | 0.56 (0.47, 0.64) | 51.2% |
| **Severe** | 0.95 (0.92, 0.98) | 92.4% | 0.58 (0.49, 0.66) | 52.7% | 0.74 (0.67, 0.81) | 70.2% |
| Incidence-based Rt was natural log-transformed. | | | | | | |
| a AUC: area under the receiver operating characteristic curve (ROC) | | | | | | |
| b Directional consistency: the proportion of days when the two estimates were simultaneously below or above 1 over the total predictable period. | | | | | | |

| **Table S5.** Distribution balance (proxied by Gini coefficient and its 95% CI) of incidence-based 𝑅𝑡 and population Ct values | | | | |
| --- | --- | --- | --- | --- |
|  | **Training sets** | | **Full waves** | |
|  | **Incidence-based Rt** | **Ct values** | **Incidence-based Rt** | **Ct values** |
| **Wave 3** | 0.185 (0.162, 0.234) | 0.154 (0.151, 0.157) | 0.4 (0.372, 0.452) | 0.158 (0.155, 0.160) |
| **Wave 4** | 0.130 (0.103, 0.171) | 0.147 (0.144, 0.150) | 0.294 (0.262, 0.333) | 0.150 (0.147, 0.152) |
| **Wave 5** | 0.448 (0.427, 0.515) | 0.156 (0.155, 0.156) | 0.457 (0.435, 0.49) | 0.162 (0.161, 0.162) |
| **Wave 6** | 0.105 (0.095, 0.123) | 0.145 (0.143, 0.146) | 0.140 (0.125, 0.156) | 0.147 (0.146, 0.148) |
| **Wave 7** | 0.173 (0.162, 0.203) | 0.141 (0.140, 0.143) | 0.130 (0.112, 0.151) | 0.143 (0.142, 0.144) |

| **Table S6.** Area under the receiver operating characteristic curve (ROC) and directional consistency between Ct-based 𝑅𝑡 and the incidence-based 𝑅𝑡 under various daily sample counts. | | | | | | | | | |
| --- | --- | --- | --- | --- | --- | --- | --- | --- | --- |
|  | **Wave 4** | | | **Wave 6** | | | **Wave 7** | | |
|  | **(Nov 2020 - Mar 2021)** | | | **(May 2022 - Sep 2022)** | | | **(Oct 2022 - Jan 2023)** | | |
|  | **Sample counts** | **AUC^a^** | **Consistency^b^** | **Sample counts** | **AUC^a^** | **Consistency^b^** | **Sample counts** | **AUC^a^** | **Consistency^b^** |
| **Training period: wave 3 (6 Jul-31 Aug 2020)** | | | | | | | | | |
| **All cases** | 151/151(100%) | 0.68 (0.6, 0.75) | 71.9% | 131/131(100%) | 0.62 (0.53, 0.7) | 67.9% | 121/121(100%) | 0.8 (0.73, 0.88) | 80.2% |
| **> 30** | 71/151(47%) | 0.72 (0.61, 0.83) | 74.6% | 116/131(89%) | 0.65 (0.56, 0.73) | 73.3% | 120/121(99%) | 0.8 (0.72, 0.87) | 80.0% |
| **> 60** | 42/151(28%) | 0.84 (0.73, 0.95) | 81.0% | 103/131(79%) | 0.7 (0.61, 0.78) | 77.7% | 119/121(99%) | 0.8 (0.72, 0.87) | 79.8% |
| **Training period: wave 4 (20 Nov - 19 Dec 2020)** | | | | | | | | | |
| **All cases** | 151/151(100%) | 0.69 (0.61, 0.76) | 73.3% | 131/131(100%) | 0.62 (0.53, 0.7) | 67.9% | 121/121(100%) | 0.81 (0.73, 0.88) | 81.0% |
| **> 30** | 71/151(47%) | 0.75 (0.64, 0.85) | 77.5% | 116/131(89%) | 0.65 (0.56, 0.73) | 73.3% | 120/121(99%) | 0.81 (0.73, 0.88) | 80.8% |
| **> 60** | 42/151(28%) | 0.86 (0.75, 0.96) | 83.3% | 103/131(79%) | 0.7 (0.61, 0.78) | 77.7% | 119/121(99%) | 0.8 (0.73, 0.88) | 80.7% |
| **Training period: wave 5 (21 Feb -22 Mar 2022)** | | | | | | | | | |
| **All cases** | 151/151(100%) | 0.67 (0.59, 0.75) | 66.4% | 131/131(100%) | 0.53 (0.49, 0.57) | 35.9% | 121/121(100%) | 0.49 (0.46, 0.53) | 37.2% |
| **> 30** | 71/151(47%) | 0.64 (0.53, 0.75) | 60.6% | 116/131(89%) | 0.54 (0.49, 0.58) | 37.9% | 120/121(99%) | 0.49 (0.45, 0.53) | 36.7% |
| **> 60** | 42/151(28%) | 0.71 (0.6, 0.83) | 64.3% | 103/131(79%) | 0.53 (0.49, 0.58) | 40.8% | 119/121(99%) | 0.5 (0.47, 0.53) | 37.0% |
| **Training period: wave 6 (28 Aug-26 Sep 2022)** | | | | | | | | | |
| **All cases** | 151/151(100%) | 0.7 (0.62, 0.77) | 74.7% | 131/131(100%) | 0.66 (0.58, 0.74) | 59.5% | 121/121(100%) | 0.53 (0.5, 0.56) | 41.3% |
| **> 30** | 71/151(47%) | 0.76 (0.66, 0.86) | 80.3% | 116/131(89%) | 0.68 (0.6, 0.77) | 63.8% | 120/121(99%) | 0.53 (0.5, 0.56) | 40.8% |
| **> 60** | 42/151(28%) | 0.91 (0.82, 1) | 90.5% | 103/131(79%) | 0.72 (0.64, 0.81) | 69.9% | 119/121(99%) | 0.53 (0.5, 0.56) | 40.3% |
| **Training period: wave 7 (18 Dec 2022-16 Jan 2023)** | | | | | | | | | |
| **All cases** | 151/151(100%) | 0.71 (0.64, 0.79) | 74.7% | 131/131(100%) | 0.66 (0.57, 0.75) | 68.7% | 121/121(100%) | 0.67 (0.59, 0.75) | 62.8% |
| **> 30** | 71/151(47%) | 0.79 (0.69, 0.89) | 80.3% | 116/131(89%) | 0.69 (0.6, 0.78) | 74.1% | 120/121(99%) | 0.67 (0.59, 0.75) | 62.5% |
| **> 60** | 42/151(28%) | 0.88 (0.77, 0.98) | 85.7% | 103/131(79%) | 0.74 (0.65, 0.83) | 79.6% | 119/121(99%) | 0.68 (0.6, 0.76) | 63.0% |
| Incidence-based Rt was natural log-transformed. | | | | | | | | | |
| a AUC: area under the receiver operating characteristic curve (ROC) | | | | | | | | | |
| b Directional consistency: the proportion of days when the two estimates were simultaneously below or above 1 over the total predictable period. | | | | | | | | | |

| **Table S7.** Numerical accuracy of model predictions using different training sets to estimate Ct-based Rt in the other four waves. | | | | | |
| --- | --- | --- | --- | --- | --- |
|  | **Wave 3** | **Wave 4** | **Wave 5** | **Wave 6** | **Wave 7** |
|  | **(July 2020 - Aug 2020)** | **(Nov 2020 - Mar 2021)** | **(Jan 2022 - May 2022)** | **(May 2022 - Sep 2022)** | **(Oct 2022 - Jan 2023)** |
| **Training period: wave 3 (6 Jul-31 Aug 2020)** | | | | | |
| **RMSE** | 0.402 | 0.553 | 0.934 | 0.416 | 0.165 |
| **MAPE** | 0.251 | 0.290 | 0.524 | 0.236 | 0.151 |
| **SMAPE** | 0.246 | 0.313 | 0.633 | 0.278 | 0.140 |
| **Training period: wave 4 (20 Nov - 19 Dec 2020)** | | | | | |
| **RMSE** | 0.470 | 0.558 | 0.963 | 0.389 | 0.170 |
| **MAPE** | 0.280 | 0.298 | 0.473 | 0.217 | 0.163 |
| **SMAPE** | 0.271 | 0.308 | 0.533 | 0.252 | 0.149 |
| **Training period: wave 5 (21 Feb -22 Mar 2022)** | | | | | |
| **RMSE** | 26.337 | 393.831 | 0.798 | 0.776 | 0.590 |
| **MAPE** | 3.454 | 16.397 | 0.673 | 0.557 | 0.544 |
| **SMAPE** | 0.720 | 0.905 | 1.084 | 0.841 | 0.770 |
| **Training period: wave 6 (28 Aug-26 Sep 2022)** | | | | | |
| **RMSE** | 0.394 | 0.585 | 0.984 | 0.469 | 0.220 |
| **MAPE** | 0.255 | 0.316 | 0.558 | 0.228 | 0.180 |
| **SMAPE** | 0.263 | 0.364 | 0.744 | 0.303 | 0.194 |
| **Training period: wave 7 (18 Dec 2022-16 Jan 2023)** | | | | | |
| **RMSE** | 0.390 | 0.719 | 0.889 | 0.454 | 0.177 |
| **MAPE** | 0.267 | 0.374 | 0.585 | 0.233 | 0.149 |
| **SMAPE** | 0.242 | 0.381 | 0.762 | 0.295 | 0.149 |
| ^*^RMSE: root mean squared error; MAPE: mean absolute percentage error; SMAPE: symmetric mean absolute percentage error. | | | | | |

**Supplementary Figures**

**Figure S1. Nowcast Ct-based 𝑅_𝑡_ during wave 4, 5, 6 and 7 in Hong Kong.** **a, b, c, d, e.** Comparing the incidence- and Ct-based 𝑅_𝑡_ over the training period (wave 3, 1 July to 31 August 2020, panel a), and the testing periods, including wave 4 (1 November 2020 to 31 March 2021, panel b), wave 5 (6 February to 22 May 2022, panel c), wave 6 (23 May to 30 September 2022, panel d) and wave 7 (1 October 2022 to 29 January 2023, panel e). **f.** The distribution of incidence-based 𝑅_𝑡_ under various intervals of Ct-based 𝑅_𝑡_ among 5 waves. Box plots indicate the interquartile ranges and medians of incidence-based 𝑅_𝑡_ under less than 0.5, 0.5 to 1, 1 to 1.5 and more than 1.5 Ct-based 𝑅_𝑡_ groups, respectively.

**Figure S2**. **Temporal distribution of population-level Ct values among mild-to-moderate and severe cases respectively during Omicron waves. a.** Temporal distribution of population-level Ct values under different severity groups. Blue lines and shaded areas indicate the average and 95% confidence intervals (CIs) of Ct values from mild-to-moderate cases estimated by a generalized additive model (GAM). Orange lines and shaded areas correspond to severe (serious/critical/fatal) cases. Black lines and shaded areas correspond to all cases. **b.** Temporal distribution of Ct skewness. Dots and vertical lines represent the mean and 95% CIs of daily Ct skewness among different severity. **c, d, e.** Correlations between the incidence-based 𝑅_𝑡_ and Ct mean and skewness during the fifth (**c**), sixth a (**d**) and sixth b (**e**) waves among different severity groups. Box plots indicate the interquartile ranges (IQR) and medians of the incidence-based 𝑅_𝑡_ under various intervals of daily Ct mean and skewness.

**Figure S3**. **Consistency of Ct-based 𝑅_𝑡_** **using the first Ct records from either mild-to-moderate or severe cases during the training wave. a.** Daily number of Ct values by date of sampling for all cases (grey bars) and for mild-to-moderate cases (blue bars) over wave 3 (n = 56 daily values). The top-right panel displayed the proportion of mild-to-moderate records over all records on each sampling date, with the red dotted line serving as the reference for the proportion of 0.7. **b-c.** Comparison of Ct-based 𝑅𝑡 estimates using all records versus records from mild-to-moderate cases (b) or severe cases (c). The mean and 95% prediction intervals of Ct-based 𝑅𝑡 estimated from the main model using all records are represented by the pink lines and shaded areas, while the mean and 95% prediction intervals of Ct-based 𝑅𝑡 estimated using mild-to-moderate records only are displayed with blue dots and vertical lines. The reference line for 𝑅𝑡 being 1 is indicated by the black dotted line.

**Figure S4**. **Consistency of Ct-based 𝑅_𝑡_** **using the first Ct records from either mild-to-moderate or severe cases during wave 4 (training wave, wave 3). a.** Daily number of Ct values by date of sampling for all cases (grey bars) and for mild-to-moderate cases (blue bars) over wave 4 (n = 127 daily values). The top-right panel displayed the proportion of mild-to-moderate records over all records on each sampling date, with the red dotted line serving as the reference for the proportion of 0.7. **b-c.** Comparison of Ct-based 𝑅𝑡 estimates using all records versus records from mild-to-moderate cases (b) or severe cases (c). The mean and 95% prediction intervals of Ct-based 𝑅𝑡 estimated from the main model using all records are represented by the pink lines and shaded areas, while the mean and 95% prediction intervals of Ct-based 𝑅𝑡 estimated using mild-to-moderate records only are displayed with blue dots and vertical lines. The reference line for 𝑅𝑡 being 1 is indicated by the black dotted line.

**Figure S5**. **Consistency of Ct-based 𝑅_𝑡_** **using the first Ct records from either mild-to-moderate or severe cases during wave 5 (training wave, wave 3). a.** Daily number of Ct values by date of sampling for all cases (grey bars) and for mild-to-moderate cases (blue bars) over wave 5 (n = 105 daily values). The top-right panel displayed the proportion of mild-to-moderate records over all records on each sampling date, with the red dotted line serving as the reference for the proportion of 0.7. **b-c.** Comparison of Ct-based 𝑅𝑡 estimates using all records versus records from mild-to-moderate cases (b) or severe cases (c). The mean and 95% prediction intervals of Ct-based 𝑅𝑡 estimated from the main model using all records are represented by the pink lines and shaded areas, while the mean and 95% prediction intervals of Ct-based 𝑅𝑡 estimated using mild-to-moderate records only are displayed with blue dots and vertical lines. The reference line for 𝑅𝑡 being 1 is indicated by the black dotted line.

**Figure S6**. **Consistency of Ct-based 𝑅_𝑡_** **using first Ct records from either mild-to-moderate or severe cases during wave 6 (training wave, wave 3). a.** Daily number of Ct values by date of sampling for all cases (grey bars) and for mild-to-moderate cases (blue bars) over wave 6a (n = 131 daily values). The top-left panel displayed the proportion of mild-to-moderate records over all records on each sampling date, with the red dotted line serving as the reference for the proportion of 0.7. **b-c.** Comparison of Ct-based 𝑅𝑡 estimates using all records versus records from mild-to-moderate cases (b) or severe cases (c). The mean and 95% prediction intervals of Ct-based 𝑅𝑡 estimated from the main model using all records are represented by the pink lines and shaded areas, while the mean and 95% prediction intervals of Ct-based 𝑅𝑡 estimated using mild-to-moderate records only are displayed with blue dots and vertical lines. The reference line for 𝑅𝑡 being 1 is indicated by the black dotted line.

**Figure S7**. **Consistency of Ct-based 𝑅_𝑡_** **using first Ct records from either mild-to-moderate or severe cases during wave 7 (training wave, wave 3). a.** Daily number of Ct values by date of sampling for all cases (grey bars) and for mild-to-moderate cases (blue bars) over wave 6b (n = 121 daily values). The top-left panel displayed the proportion of mild-to-moderate records over all records on each sampling date, with the red dotted line serving as the reference for the proportion of 0.7. **b-c.** Comparison of Ct-based 𝑅𝑡 estimates using all records versus records from mild-to-moderate cases (b) or severe cases (c). The mean and 95% prediction intervals of Ct-based 𝑅𝑡 estimated from the main model using all records are represented by the pink lines and shaded areas, while the mean and 95% prediction intervals of Ct-based 𝑅𝑡 estimated using mild-to-moderate records only are displayed with blue dots and vertical lines. The reference line for 𝑅𝑡 being 1 is indicated by the black dotted line.

**Figure S8**. **Daily distributions of population-level Ct values and delays from onset to sampling by sampling date. a-c.** Daily distributions of Ct values by sampling date for all cases (panel a, n = 120768 Ct records), mild-to-moderate cases (panel b, n = 77647 Ct records) and severe cases (panel c, n = 43121 Ct records). Orange lines and shaded areas depict the daily mean and 95% CIs of Ct values estimated from Generalized Additive Models (GAM). Black dotted line indicates the reference of Ct being 35. **d-f.** Daily distributions of delays from onset to sampling by sampling date for all cases (panel d, n = 48599 onset- to-sampling delays), mild-to-moderate cases (panel e, n = 31600 onset-to-sampling delays) and severe cases (panel f, n = 16999 onset-to-sampling delays). Black lines and shaded areas represent the daily mean and 95% CIs of delays smoothed by GAM. Dark grey vertical lines and dots (deep orange for daily Ct values in panels a-c and deep green for daily delays in panels d-f) illustrate the interquartile range (IQR) and median of daily values, while light grey vertical lines denote the minimum and maximum values observed on each day. Black dotted lines indicate the reference of delay being 3 and 12, respectively.

**Figure S9**. **Daily distributions of delays from the initial sampling to clinical outcomes.** Daily distributions of delays from the initial sampling of Ct values to clinical outcomes by clinical outcomes date for severe cases (n = 33839 onset- to-sampling delays). Black lines and shaded areas represent the daily mean and 95% CIs of delays smoothed by GAM. Dark grey vertical lines and dots (deep blue for daily delays) illustrate the interquartile range (IQR) and median of daily values, while light grey vertical lines denote the minimum and maximum values observed on each day. Black dotted line indicates the reference of delay being 2.
