## Supplementary figures and images for "Effective real-time transmission estimations incorporating population viral load distributions amid SARS-CoV-2 variants and pre-existing immunity"

### Fig_S1.pdf

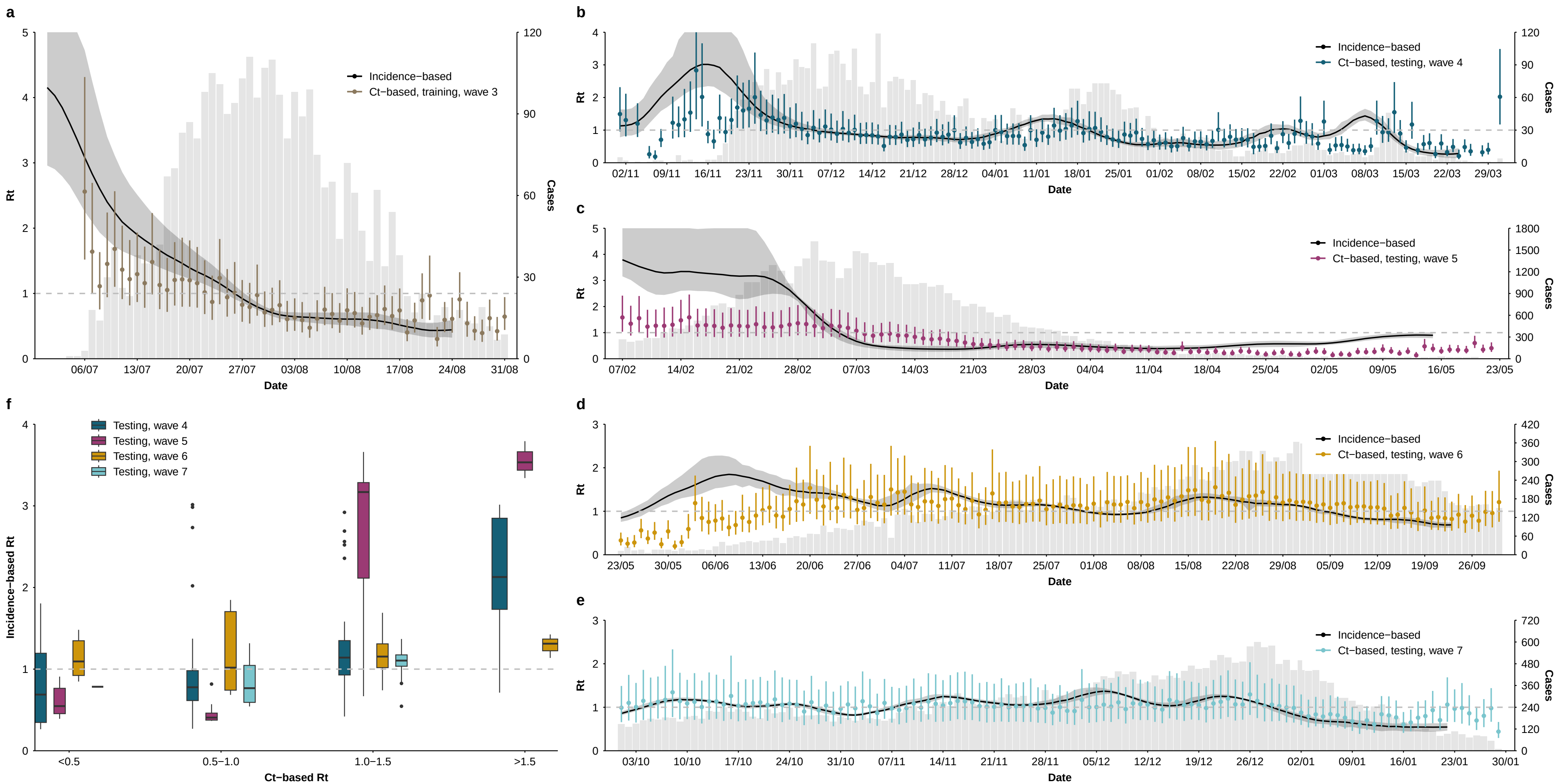

### Fig_S2.pdf

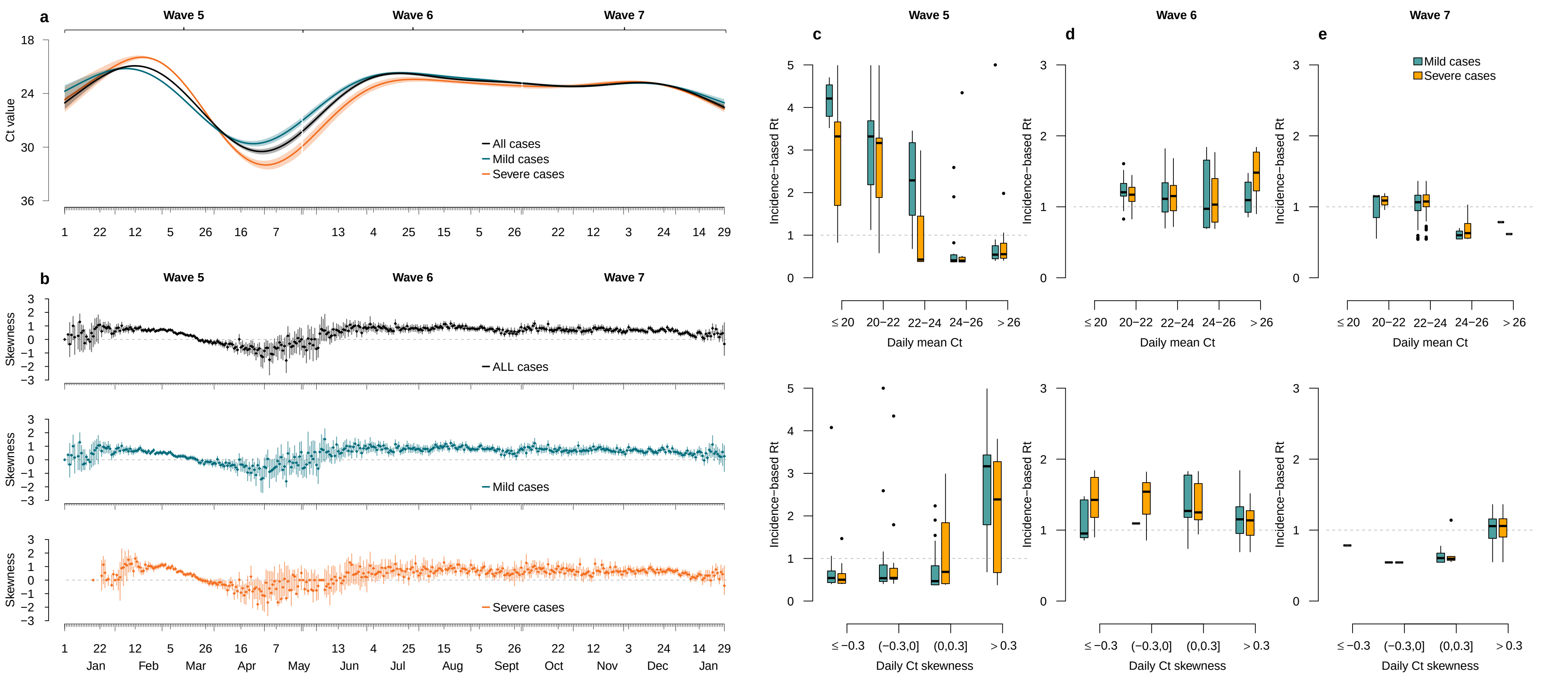

### Fig_S3.pdf

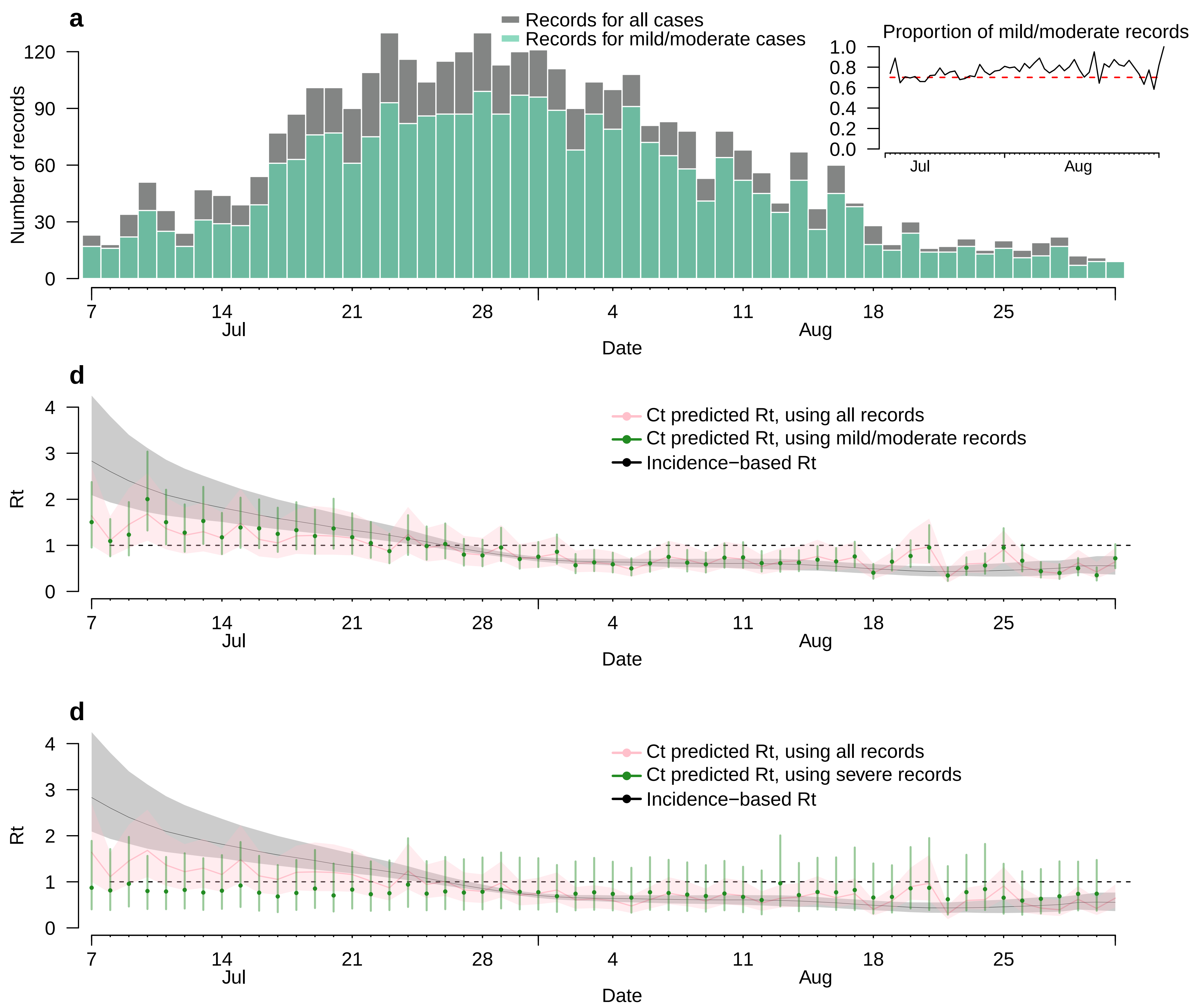

### Fig_S4.pdf

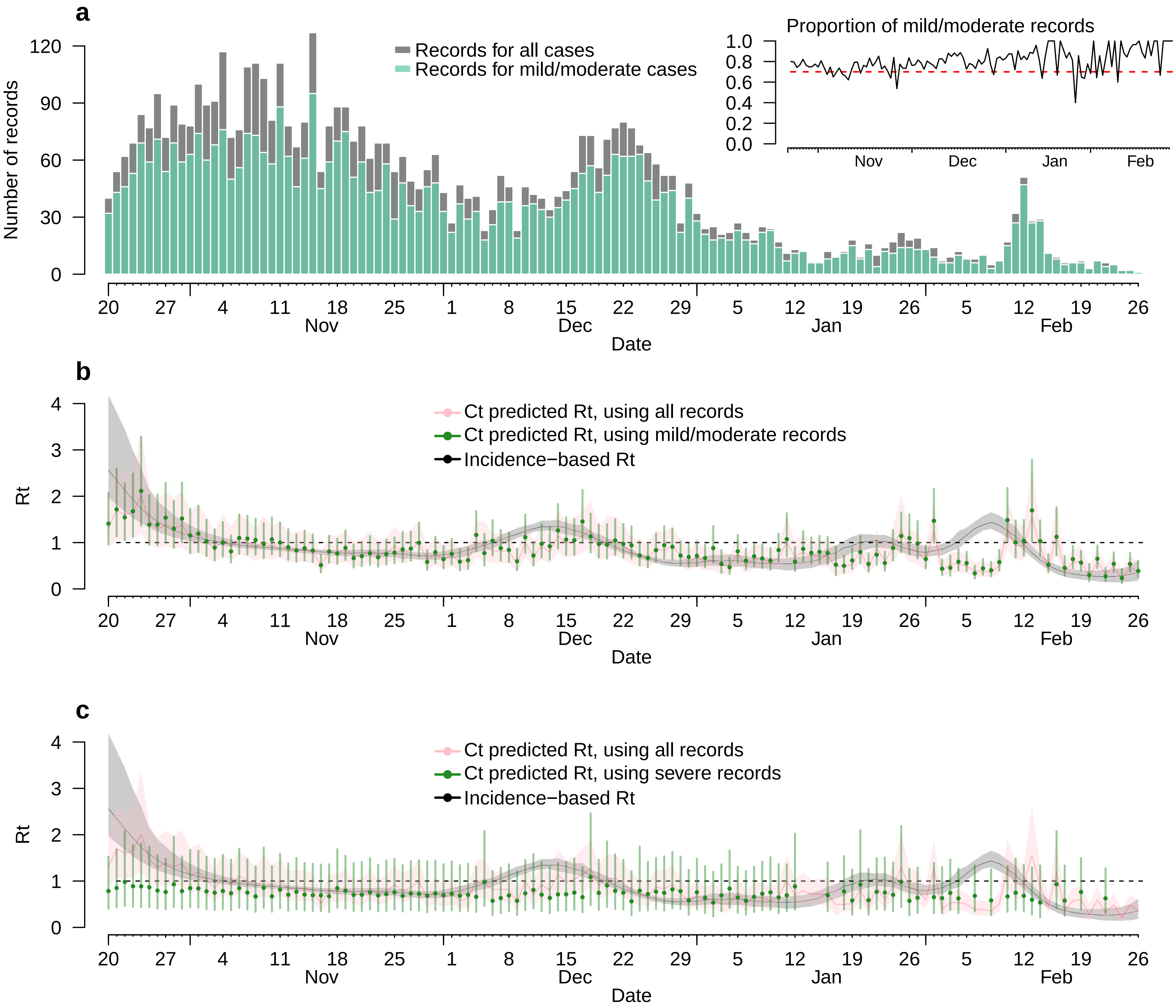

### Fig_S5.pdf

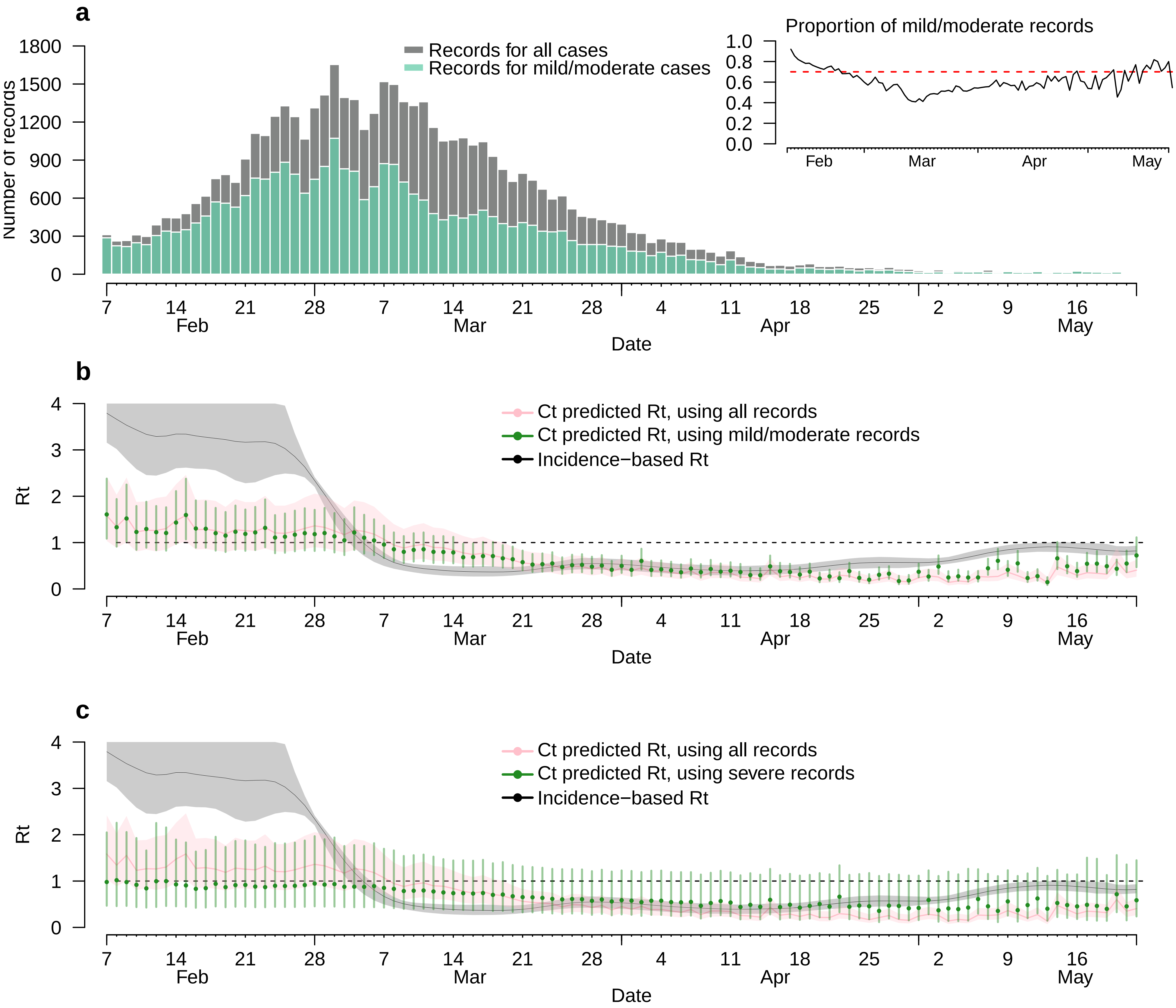

### Fig_S6.pdf

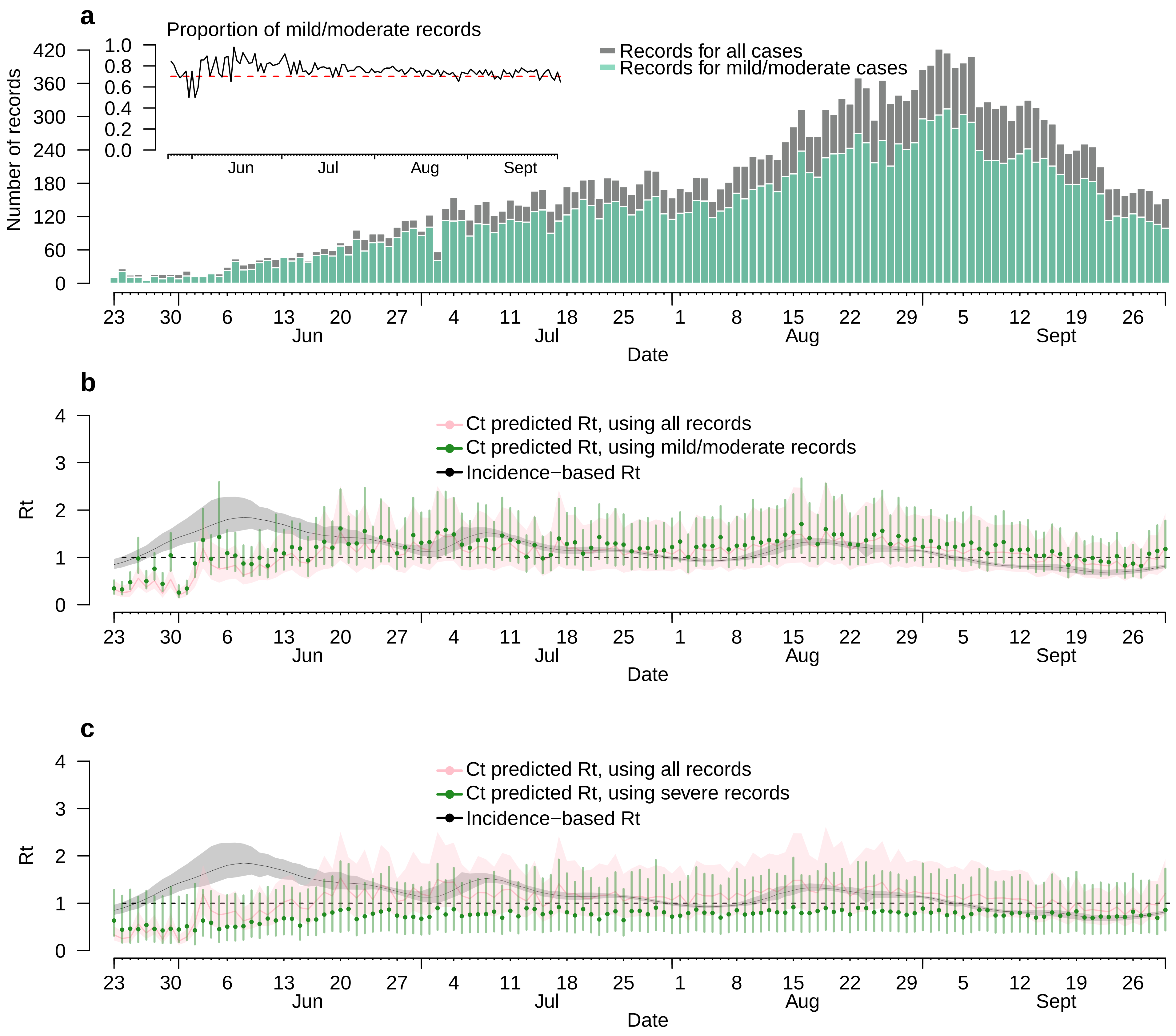

### Fig_S7.pdf

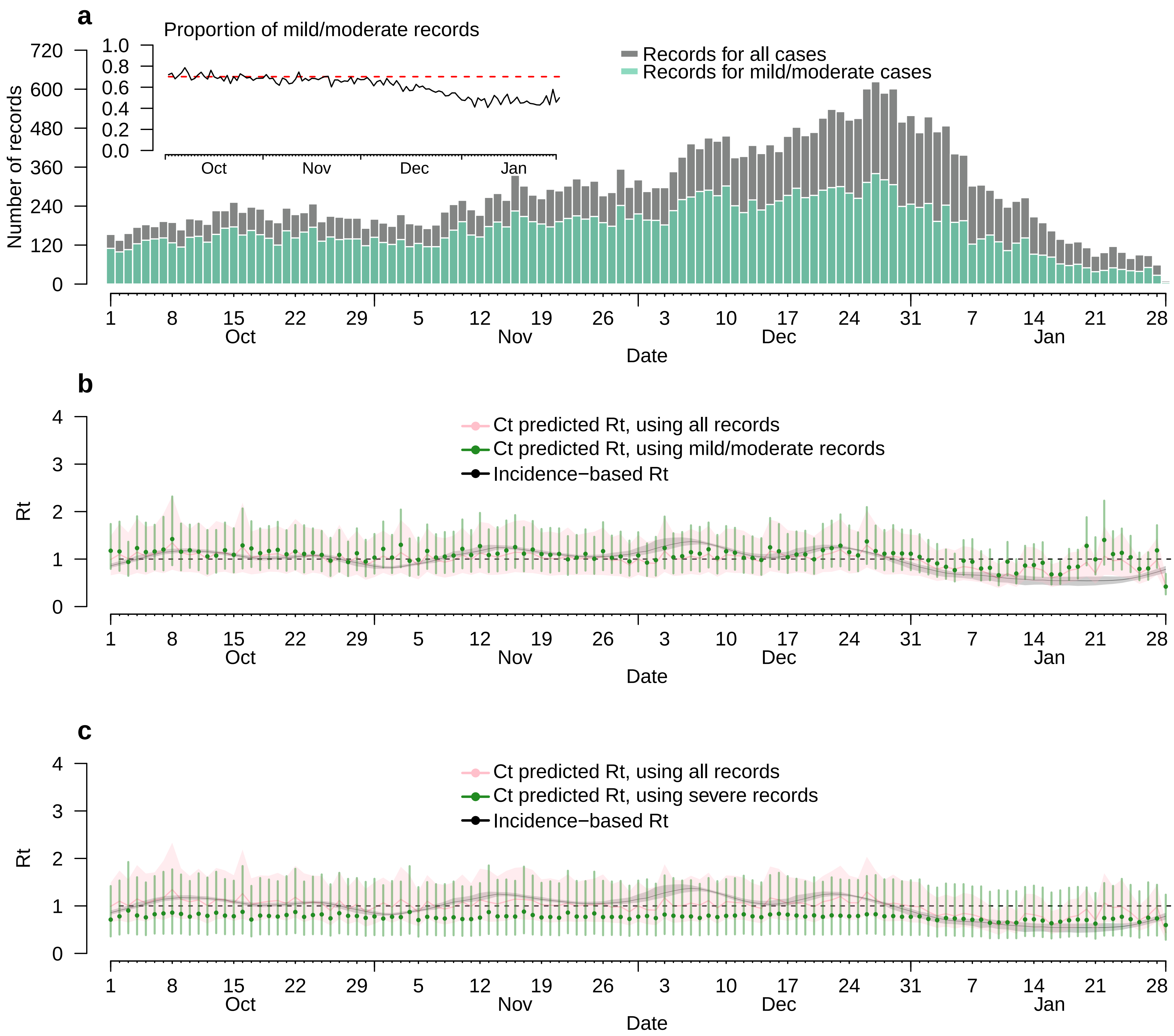

### Fig_S8.pdf

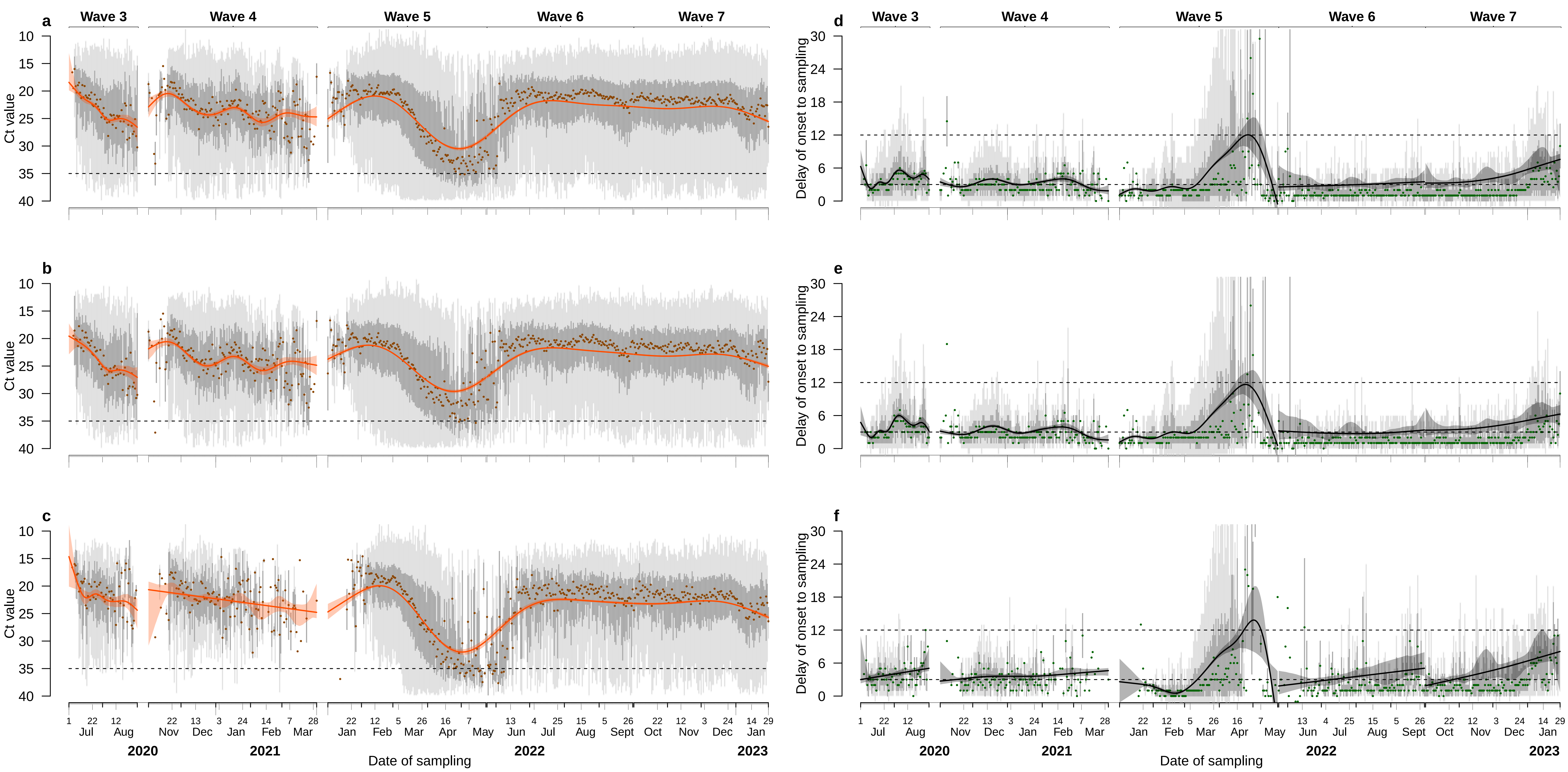

### Fig_S9.pdf

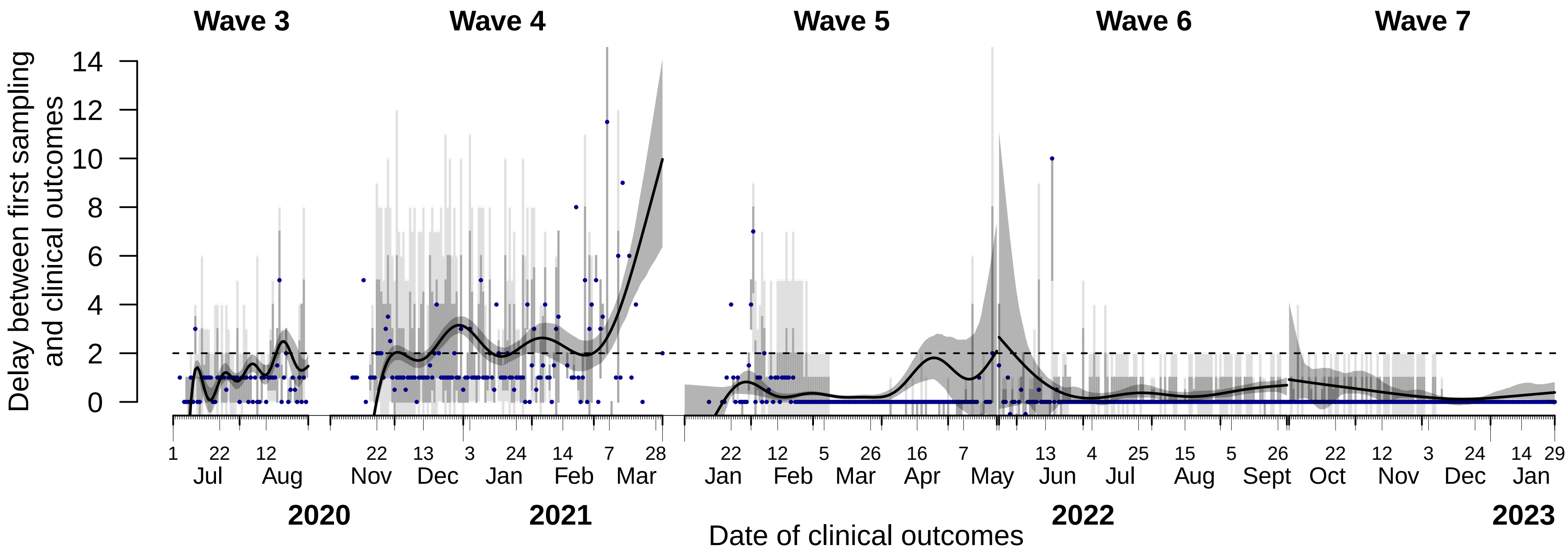
